# Pilot trial of perampanel on peritumoral hyperexcitability and clinical outcomes in newly diagnosed high-grade glioma

**DOI:** 10.1101/2024.04.11.24305666

**Authors:** Steven Tobochnik, Michael S. Regan, Maria K. C. Dorotan, Dustine Reich, Emily Lapinskas, Md Amin Hossain, Sylwia Stopka, Sandro Santagata, Melissa M. Murphy, Omar Arnaout, Wenya Linda Bi, E. Antonio Chiocca, Alexandra J. Golby, Michael A. Mooney, Timothy R. Smith, Keith L. Ligon, Patrick Y. Wen, Nathalie Y. R. Agar, Jong Woo Lee

## Abstract

**Background:** Glutamatergic neuron-glioma synaptogenesis and peritumoral hyperexcitability promote glioma growth in a positive feedback loop. The objective of this study was to evaluate the feasibility and estimated effect sizes of the AMPA-R antagonist, perampanel, on intraoperative electrophysiologic hyperexcitability and clinical outcomes.

**Methods:** An open-label trial was performed comparing perampanel to standard of care (SOC) in patients undergoing resection of newly-diagnosed radiologic high-grade glioma. Perampanel was administered as a pre-operative loading dose followed by maintenance therapy until progressive disease or up to 12-months. SOC treatment involved levetiracetam for 7-days or as clinically indicated. The primary outcome of hyperexcitability was defined by intra-operative electrocorticography high frequency oscillation (HFO) rates. Seizure-freedom and overall survival (OS) were estimated by the Kaplan-Meier method. Tissue concentrations of perampanel, levetiracetam, and metabolites were measured by mass spectrometry.

**Results:** HFO rates were similar between perampanel-treated and SOC cohorts. The trial was terminated early after interim analysis for futility, and outcomes assessed in 11 patients (7 perampanel-treated, 4 SOC). Over a median 281 days of post-enrollment follow-up, 27% of patients had seizures, including 14% treated with perampanel and 50% treated with SOC. OS in perampanel-treated patients was similar to a glioblastoma reference cohort (p=0.81). Glutamate concentrations in surface biopsies were positively correlated with HFO rates in adjacent electrode contacts and were not significantly associated with treatment assignment or drug concentrations.

**Conclusions:** A peri-operative loading regimen of perampanel was safe and well-tolerated, with similar peritumoral hyperexcitability as in levetiracetam-treated patients. Maintenance anti-glutamatergic therapy was not observed to impact survival outcomes.

## INTRODUCTION

Diffuse gliomas are commonly associated with seizures over the course of disease.^1^ The detection of peritumoral hyperexcitability by electrophysiologic brain recording techniques serves as the primary biomarker of tumor-related epileptogenesis. Although multiple mechanisms of tumor-related epileptogenesis exist, a role for increased glutamatergic signaling in the glioma microenvironment appears to be a critical early event in promoting hyperexcitability,^2^ mediated in part by glioma cell-autonomous release of glutamate and impaired re-uptake from the extracellular space.^3^ Recent evidence also indicates a bidirectional interaction between tumor growth and neuronal hyperexcitability driven by the development of functional glutamatergic synapses between neurons and glioma cells, and increased AMPA receptor (AMPA-R) activation.^4–6^ In preclinical studies, excessive neuronal activity drives the feedforward release of neuroligin-3, leading to induction of PI3K-mTOR activity and glioma cellular proliferation.^7^

These findings have led to a renewed interest in the potential for anti-seizure medications (ASMs) targeting hyperexcitability to improve tumor outcomes as well as seizure control in glioma-related epilepsy. However, post hoc analyses in neuro-oncology cohorts evaluating tumor outcomes associated with ASM exposure have provided conflicting results and have been inherently limited by indication bias.^8–19^ The selective post-synaptic AMPA-R antagonists, talampanal and perampanel, have demonstrated promising anti-seizure effects,^20–24^ although their impact on clinical tumor outcomes remains uncertain.^17–19^ Furthermore, it is unclear whether targeting glutamatergic signaling provides an advantage over targeting other mechanisms of excessive synaptic activity with first-line broad-spectrum ASMs such as levetiracetam. Currently, standard of care involves the use of levetiracetam indefinitely for treatment of clinical seizures or for about 1 week post-operatively for seizure prophylaxis, with other ASMs generally used when levetiracetam is ineffective or poorly tolerated.

High gamma activity, reflective of synchronized neuronal firing at peritumoral margins,^25,26^ may be a promising intra-operative biomarker of hyperexcitability for the evaluation of targeted therapies. In this pilot trial of patients with newly diagnosed radiologic high-grade glioma (HGG), we assessed the feasibility and effects of a peri-operative loading regimen of perampanel on peritumoral high frequency oscillations (HFOs) using intra-operative electrocorticography (ECoG) and correlative tissue biomarkers, followed by long-term maintenance therapy to estimate clinical effect sizes for seizure and survival outcomes.

## MATERIALS AND METHODS

### Study Design

This was a single-site open-label non-randomized prospective trial (ClinicalTrials.gov Registration: NCT04497142). Patients were screened and enrolled at Brigham and Women’s Hospital and Dana-Farber Cancer Institute (DFCI). The primary outcome was the rate of peritumoral HFOs by intra-operative ECoG. Secondary outcomes included time to first post-treatment seizure, overall survival (OS), and tissue glutamate concentration by mass spectrometry imaging (MSI). The study was approved by the Institutional Review Board of Dana-Farber Cancer Institute (#20-059).

### Eligibility Criteria

Inclusion criteria were adults aged 18 years or older with radiologic evidence of newly-diagnosed HGG planned for resection, capacity to provide informed consent, Karnofsky performance status ≥60, and adequate organ and marrow function (leukocytes ≥3,000/mcL, absolute neutrophil count ≥1,500/mcL, platelets ≥100,000/mcL, total bilirubin ≤ institutional upper limit of normal (ULN), AST/ALT ≤3x ULN, GFR ≥30 mL/min/1.73m^2^). Exclusion criteria included participants with concurrent malignancy, active suicidal ideation (Columbia Suicide Severity Rating Scale type 4-5), active treatment with >1 ASM, or tumor-associated seizures >1 month prior to resection.

### Treatment Protocol

Eligible participants were consented to receive either perampanel treatment (Cohort 1) or standard of care (Cohort 2). Due to the short time window for enrollment, patients were informed of both study arms and given the option to enroll in either arm in a non-randomized manner. Participants in Cohort 1 received a loading dose of perampanel 6 mg at 12-36 hours (0.11-0.34 half-lives) before surgery and were continued on a maintenance dose of perampanel 4 mg daily until progressive disease (PD) by Response Assessment in Neuro-Oncology (RANO) criteria or up to 12 months,^27^ allowing for dose adjustments as clinically indicated. Those who were already treated with levetiracetam pre-operatively received their last dose ≥36 hours (about 5 half-lives) before surgery and were transitioned to perampanel. Participants in Cohort 2 were treated as clinically indicated according to institutional standards, involving an intra-operative loading dose of levetiracetam followed by levetiracetam 1000-1500 mg/day for 7 days post-operatively in the absence of prior seizures, and continued indefinitely at the discretion of the treating physician in patients with a history of seizures. All participants received standard of care tumor-targeted treatment. Co-enrollment in a clinical trial with or without additional experimental therapies was allowed.

### Safety Assessments

Adverse effects were graded according to the revised NCI Common Terminology Criteria for Adverse Events (CTCAE) version 5.0.

### Electrocorticography (ECoG)

Intraoperative ECoG was performed using an XLTEK system (Natus Medical Inc, Pleasanton, CA), sampled at 1024 Hz per channel with a subdural 20-contact electrode grid overlying the tumor margins. The grid was trimmed as needed to fit the craniotomy. At least 10 minutes prior to ECoG, inhaled anesthetics and propofol infusion were stopped as per standard institutional practices. Recording was performed for 10 minutes in each case except for one in which recording was performed for 2 minutes due to excessive cerebral edema.

### High Frequency Analysis

High frequency oscillations (HFOs) were evaluated post-operatively. ECoG time series were analyzed after common average referencing. Channels with excessive artifact were excluded from the average reference and HFO analysis, and channels with continuous spiking activity were excluded from the average reference but retained for HFO analysis. HFO events in the 80-500 Hz band were detected using the Hilbert transform (threshold 3 standard deviations) in an automated manner,^28^ followed by visual review to exclude false-positive detections related to electrical or filtering artifacts. HFO events were required to have at least 4 consecutive oscillations distinct from the background, as previously described.^29^ HFO rates were calculated per channel per minute.

### Imaging Analysis

Pre-operative contrast-enhanced MRIs were coregistered to ECoG electrode contacts and biopsy sites in patient space (Figure 1a) using the intra-operative neuronavigation system and 3D Slicer (slicer.org).^30^ Brainlab was used in 11 patients and Medtronic Stealth in 1 patient. Site localization was defined as peritumoral if ≤1 cm of contrast enhancing tumor in any dimension, in cortex if >1 cm from contrast enhancing tumor in each dimension, and in tumor if exclusively overlying contrast enhancing tumor for >1 cm in each dimension.

**Figure 1.**
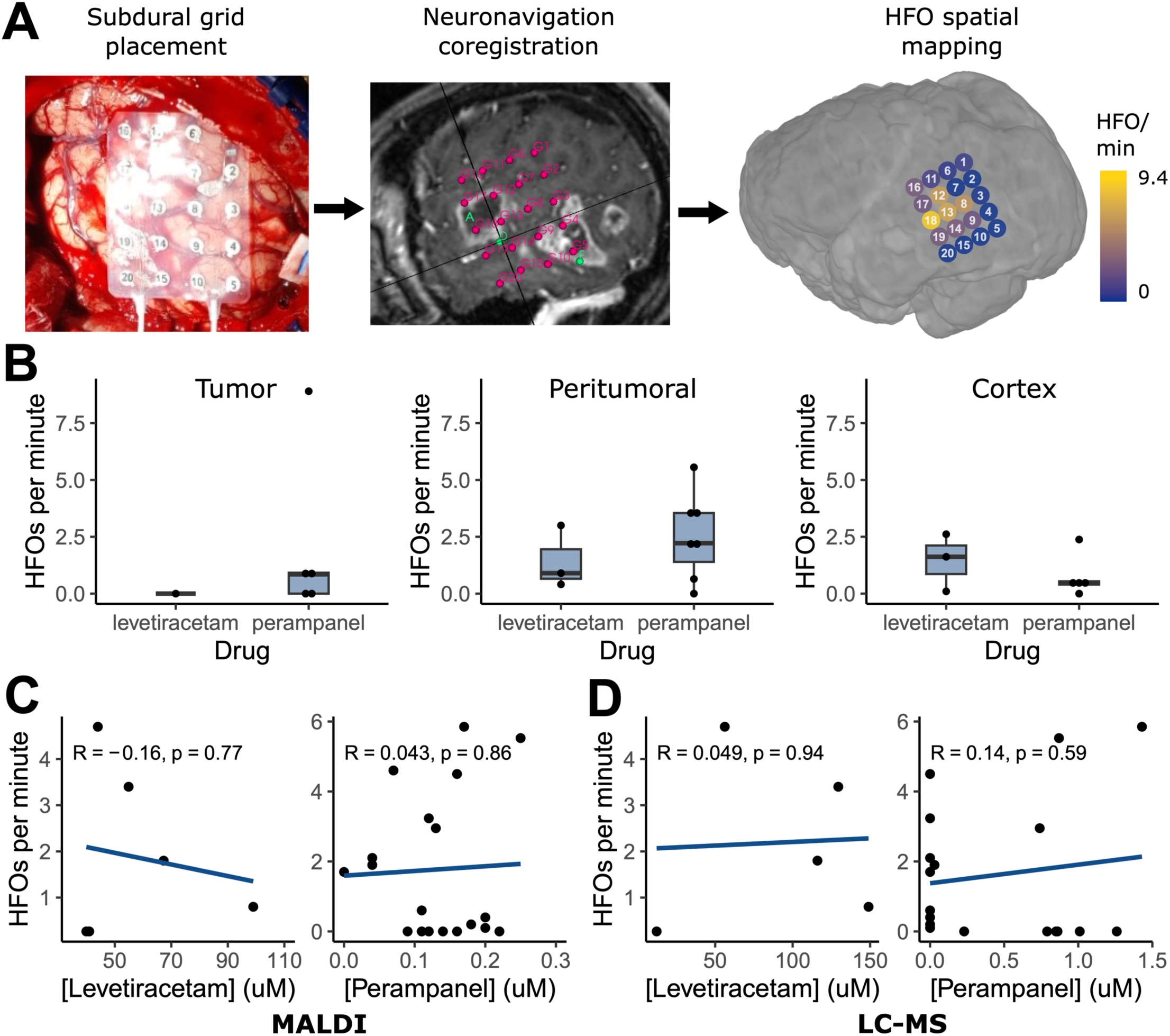
Peritumoral hyperexcitability analysis. (A) Workflow for spatial mapping of HFOs and biopsy sites to patient space. (B) HFO rates between perampanel and levetiracetam treated patients at MRI defined locations relative to the contrast-enhancing tumor margin. All comparisons were non-significant. (C,D) Correlations between HFO rates and tissue drug concentration at adjacent biopsy sites measured by MALDI (C) and LC-MS (D).

### Tissue Biomarker Analysis

Multiple tissue biopsies were obtained for each patient during standard of care maximal safe resection. Surface biopsies from within 1 cm of the ECoG grid were analyzed for associations between tissue biomarkers and HFO activity. All biopsies with sufficient tissue were analyzed for ASM concentration (levetiracetam and perampanel as applicable), glutamate concentration, and lactate/pyruvate ratio using mass spectrometry. Matrix-assisted laser desorption/ionization (MALDI) mass spectrometry imaging was performed for spatially resolved analysis in *ex vivo* tissue slices, and total concentrations for each slice validated using liquid chromatography-tandem mass spectrometry (LC-MS).

#### MALDI Tissue Preparation

Tissue specimens were snap frozen on dry ice in 1.5 mL cryovials, mounted onto a specimen chuck, and sectioned at 10 µm thickness (CM1950, Leica Biosystems Teaneck, NJ). The tissue sections were thaw-mounted onto indium tin oxide (ITO) slides (576352, Sigma-Aldrich, St. louis, MO). Mimetics for MALDI quantitation were performed using a tissue microarray (TMA) mold.^31^ 15N-glutamate (332143, Sigma-Aldrich) was resuspended and pipetted into rat tail collagen I in microcentrifuge tubes. 15N-glutamate concentrations ranging from 0 – 30 mM were pipetted into 1.5 mm core diameter channels of the 40% gelatin TMA mold. Perampanel and levetiracetam were mixed with homogenized human brain tissue for final concentrations ranging from 0 – 25 μM and 0 – 300 μM, respectively. The microcentrifuge tubes with spiked tissue were frozen and punched into the 1.5 mm core diameter TMA. Mimetics were sectioned as tissue specimens and thaw mounted onto the same slide for MALDI acquisition. Serial sections of tissue specimens were mounted on glass microscope slides for hematoxylin and eosin (H&E) staining. ITO slides with tissue and mimetic mold sections were placed in a vacuum desiccator for 15 minutes prior to matrix application. For glutamate quantitation, 1,5 - Diaminonaphthalene (DAN) (4.38 mg/mL) matrix was dissolved in 4 mL HPLC water, 0.5 mL HCl, and 4.5mL HPLC ethanol. D5 – glutamate (34840, Cayman Chemicals, Ann Arbor MI) was added to matrix solution as an internal standard for final concentration of 250 μM. DAN-HCl and IS were applied to the ITO slide using M5 TM Sprayer (HTX Technologies, Chapel Hill, NC) at a flow rate of (0.09 mL/min), spray nozzle velocity (1200 mm/min), nitrogen gas pressure (10 psi), nozzle spray temperature (75 °C), track spacing (2mm), heated tray (55 °C), for two total passes. For perampanel and levetiracetam quantitation, 2,5-dihydroxybenzoic acid (160 mg/mL) matrix was dissolved in 70:30 methanol: 0.1% TFA with 1 % DMSO and applied using the M5 TM sprayer at a flow rate (0.18 mL/min), spray nozzle velocity (1200 mm/min), nitrogen gas pressure (10 psi), spray nozzle temperature (75 °C), and track spacing (2 mm) for four passes. Optical microscopy images of the H&E-stained serial tissue sections were acquired using a 10x objective (Zeiss Observer Z.1, Oberkochen, Germany).

#### MALDI Mass Spectrometry Imaging

A multiple reaction monitoring (MRM) method was used for quantitative imaging of perampanel and levetiracetam by monitoring the transition of the precursor ion fragment to product ion (Perampanel: *m/z* 350.13 → 219.09, Levetiracetam: *m/z* 171.1 → 126.09) using a dual source trapped ion-mobility spectrometry time-of-flight (timsTOF) fleX mass spectrometer (Bruker Scientific LLC, Billerica, MA) in positive ion mode. Data was acquired between *m/z* 100-650. Instrument parameters were optimized using ESI with an infusion of either perampanel or levetiracetam to adjust the ion transfer funnels, quadrupole, collision cell, and focus pre-TOF parameters, and an Agilent tune mix solution (Agilent Technologies, Santa Clara, CA) was used to calibrate the mass range. Tandem MS parameters were set for a collision energy of 15 eV for perampanel and 9 eV for levetiracetam with an isolation width of 3 m/z for both compounds. MALDI MS images were acquired with a laser repetition set to 10,000 Hz with 1,000 laser shots per 50 µm pixel. Quantitative imaging of glutamate was acquired on a 15 Tesla solariX FTICR mass spectrometer with a dual ESI/MALDI source (Bruker Daltonics, Billerica, MA) operating in negative-ion mode. The MSI methods were optimized and calibrated by using the electrospray source (Supplementary Figure 1). Agilent tune mix solution was used to calibrate the mass range. The data acquisition parameters were optimized to a defined pixel step size of 100 μm covering the *m/z* range of 92.15–3000. Each pixel consisted of 250 laser shots with a frequency of 1000 Hz.

#### MALDI Data Processing

SCiLS Lab software (version 2024a premium, Bruker Scientific LLC, Billerica, MA) was used for data visualization and normalization to internal standards. The average ion intensity for each spiked TMA sectioned core area was plotted against corresponding analyte concentrations resulting in a limit of detection (LOD) of 0.03 µM and a limit of quantification (LOQ) of 0.12 µM for perampanel, LOD of 0.21 µM and LOQ of 0.71 µM for levetiracetam, and a LOD of 0.0005 µM and LOQ of 0.0017 µM for glutamate.

### Drug Quantitation by LC-MS

Tissue specimens were homogenized and diluted 10-fold using LC-MS grade water before extraction using acetonitrile (1:1 [v/v], 0.2 µM of chlorpropamide as an internal standard with acetonitrile) according to previously published methods.^31,32^ Both perampanel and levetiracetam were quantified using a 5500 ESI triple-quadruple mass spectrometer (Sciex, Framingham, MA) coupled with an Acquity ultra pressure liquid chromatograph (UPLC) (Waters Corp., Milford, MA) as previously described.^33,34^ The instrument was operated in positive-ion mode and 5 μL of each sample was injected on an Acquity HSS T3, 1.8 μM, 2.1 mm × 150 mm UPLC column (Milford, MA) at 50 °C. Solvent A (water with 0.1% FA) and solvent B (acetonitrile with 0.1% FA) were combined in a 5-min gradient: 0−0.45 min, 10% B; 3.00−3.75 min, 95% B; 3.80−5.00 min, 10% B. The analyst software v1.7.3 (Framingham, MA) was used to control the instrument and to perform data analysis. All the samples were injected in duplicate and quantified against a twelve-point standard curve with a 5000-fold dynamic range (0.0005 μM to 2.5 μM). The ESI mass spectrometer was operated in MRM mode with the following MRM transitions as precursor to production ions: perampanel, *m/z* 350.02 → 247.18; levetiracetam, *m/z* 171.00 → 126.06; chlorpropamide, *m/z* 276.92 → 110.95 along with declustering potential (DP) of 80 V, 25 V, and 46 V; collision energy (CE) 40 V, 21 V, and 47 V, respectively. Curtain gas was maintained at (CUR) 20 along with ion gas source 50, ion spray voltage 5000, and temperature 600 °C.

### Statistical Analysis

Associations between categorical and continuous data were tested using Fisher’s Exact and Mann-Whitney U tests, respectively. Tissue mass spectrometry data were analyzed using fixed-effects and mixed-effects linear regression with p-values estimated by parametric bootstrapping. Survival and seizure-free probabilities were estimated using the Kaplan-Meier method. OS was compared between patients treated with perampanel in Cohort 1 and a reference cohort of patients with an integrated pathologic diagnosis of glioblastoma by 2021 WHO classification, treated between 2013-2022, extracted from the DFCI Oncology Data Retrieval System (OncDRS) database.^35^ Right-censored OS times from diagnosis were compared between groups using the log-rank test. All tests were two-sided with a significance level of 0.05. Statistical analyses were performed using R 4.2.2 (The R Foundation).^36^

## RESULTS

### Cohort Characteristics

There were 12 patients enrolled, including eight on perampanel protocol (Cohort 1) and four on standard of care protocol (Cohort 2). One participant in Cohort 1 became ineligible after pathology revealed metastatic adenocarcinoma and was removed from further analysis. Clinical and demographic characteristics are shown in Table 1.

**Table 1.** Participant clinical characteristics.

|  | Total<br>N=11 | Cohort 1 (PER)<br>N=7 | Cohort 2 (LEV)<br>N=4 |
| --- | --- | --- | --- |
| Age, years, mean (range) | 62 (37-71) | 64 (50-71) | 57 (37-69) |
| Sex, female, N (%) | 6 (55%) | 3 (43%) | 3 (75%) |
| Integrated pathology, N (%) |  |  |  |
| Glioblastoma | 10 (91%) | 7 (100%) | 3 (75%) |
| Astrocytoma, IDH-mutated | 1 (9%) | 0 (0%) | 1 (25%) |
| WHO grade, N (%) |  |  |  |
| 4 | 10 (91%) | 7 (100%) | 3 (75%) |
| 3 | 1 (9%) | 0 (0%) | 1 (25%) |
| KPS, median (range) | 80 (70-100) | 80 (70-90) | 95 (80-100) |
| Gross total resection, N (%) | 6 (55%) | 2 (29%) | 4 (100%) |
| Tumor localization, N (%)<sup>a</sup> |  |  |  |
| Frontal | 3 (27%) | 2 (29%) | 1 (25%) |
| Temporal | 8 (73%) | 6 (86%) | 2 (50%) |
| Parietal | 3 (27%) | 1 (14%) | 1 (25%) |
| Pre-operative seizures, N (%) | 4 (36%) | 4 (57%) | 0 (0%) |
| Intra-operative seizures, N (%) | 1 (9%) | 0 (0%) | 1 (25%) |
| Post-operative seizures, N (%) | 2 (18%) | 1 (14%) | 1 (25%) |

<sup>a</sup> Each affected lobe counted in multilobar cases
IDH, Isocitrate dehydrogenase; KPS, Karnofsky performance status; LEV, levetiracetam; PER, perampanel; WHO, World Health Organization

All patients in Cohort 1 received perampanel 6 mg the day prior to surgery, 4 mg the morning prior to surgery, and continued 4 mg daily post-operatively. No adverse effects occurred pre-operatively or within the first post-operative week with this loading protocol. All patients in Cohort 2 received levetiracetam 500mg intravenously at the time of craniotomy. During active follow-up, there were no unanticipated or serious adverse events probably/definitely attributed to study interventions in either cohort.

There were 42 distinct tissue specimens from 11 cases included in analysis, with a median of 4 per patient (range 2-5). These included 27 biopsies (median 3, range 1-4 per patient) from cortical sites that were correlated with electrocorticography data and 15 biopsies (median 1, range 0-3 per patient) from deeper sites.

### Intra-operative High Frequency Oscillations

Peritumoral hyperexcitability was quantified by measuring the rate of 80-500 Hz HFOs per minute in each channel by intra-operative ECoG. HFOs were detected in 10/11 (91%) cases including a total of 171 ECoG recording sites. HFOs occurred at a rate of at least 1.0/minute at 60 (35%) sites, were absent at 76 (44%) sites, and demonstrated borderline rates between 0.0-1.0/minute at the remaining 35 (20%) sites. Compared to sites overlying tumor, HFO rates were higher at peritumoral sites (mean 2.1 vs 1.2 per minute, p=0.016) and normal appearing cortex (mean 1.4 vs 1.2 per minute, p=0.058).

Mean HFO rates were similar between Cohort 1 and Cohort 2 (Figure 1b) in normal appearing cortex (0.8 vs 1.4, p=0.57), peritumoral margins (2.5 vs 1.4, p=0.52), and overlying tumor (2.1 vs 0.0, p=0.53). The trial was terminated early after a planned interim analysis was conducted in the initial 10 cases after excluding one patient with non-glial pathology. A total of 159 ECoG contact sites from these 10 patients (7 treated with perampanel, 3 treated with levetiracetam) were included. Based on the mean HFO rates at 50% of target accrual, the conditional probability of rejecting the null hypothesis at 100% accrual was 4.6%.

### Seizure-free Survival

Over a median 281 (range 87-364) days in active follow-up, 3/11 (27%) patients had seizures while on study protocol (Figure 2a), including 1/7 (14%) treated with perampanel (Cohort 1) and 2/4 (50%) treated with standard of care (Cohort 2). Among these three, pre-operative seizures were present in the patient treated with perampanel, but not in either patient in Cohort 2 treated with peri-operative levetiracetam. Of the two patients with seizures in Cohort 2, one had exclusively intra-operative bilateral tonic-clonic seizures and was later transitioned to lacosamide due to adverse effects of levetiracetam, and the other completed a 7-day post-operative course of levetiracetam with a first focal impaired awareness seizure occurring 226 days after tumor resection with stable disease.

**Figure 2.**
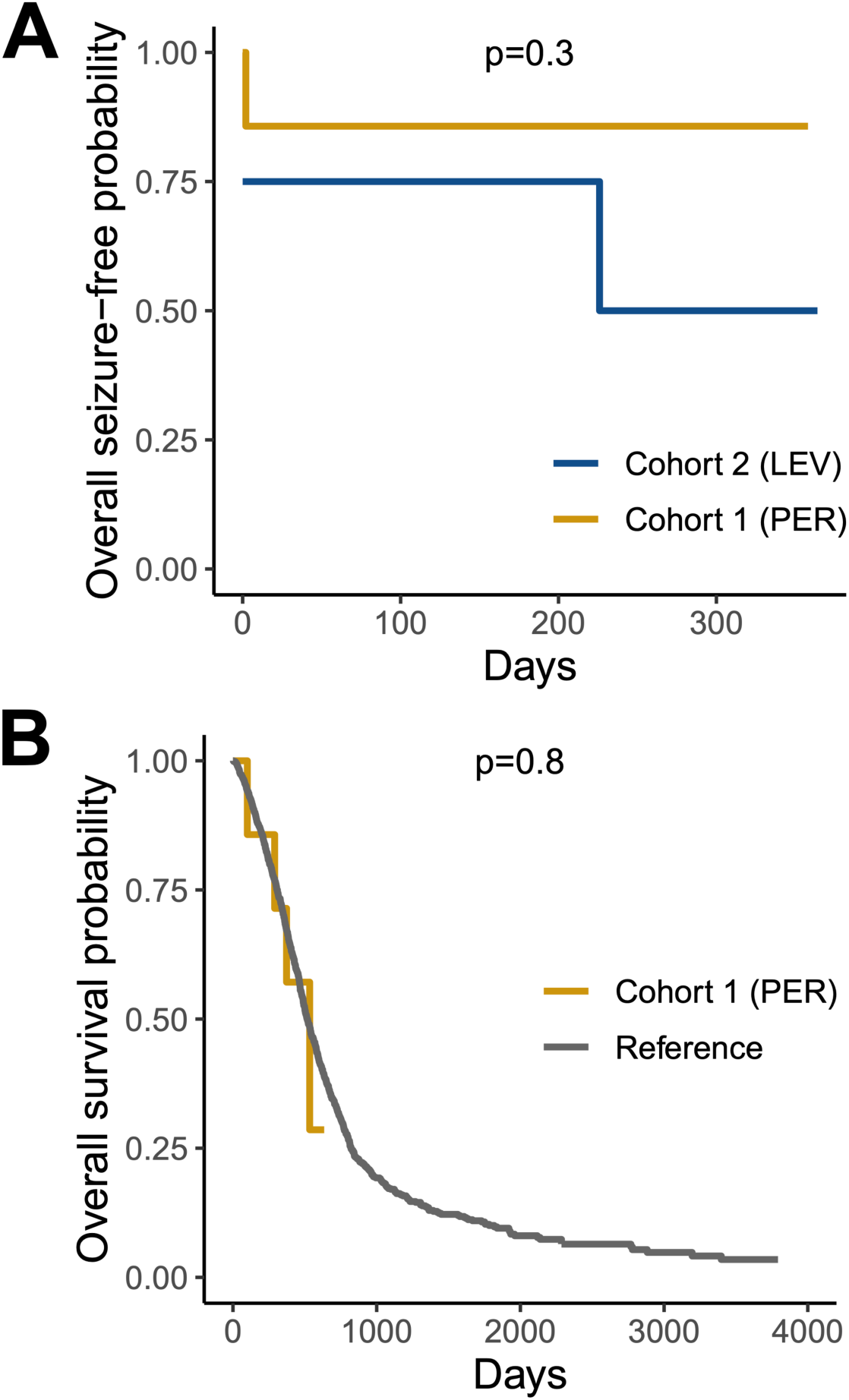
Clinical outcomes survival analyses. (A) Kaplan-Meier curve for seizure-free survival in Cohort 1 (PER) versus Cohort 2 (LEV). (B) Kaplan-Meier curve for overall survival in Cohort 1 (PER) versus a reference glioblastoma cohort.

### Overall Survival

An adult DFCI glioblastoma reference cohort (N=1445, 39.8% female, mean age 60.8 [range 21-94] years) was used as a comparator to estimate the effect size of long-term maintenance perampanel treatment on OS. OS in patients treated with perampanel in Cohort 1 (median 534 days, 95%CI 290-NA) was similar to this reference cohort (median 516 days, 95%CI 492-545, p=0.81, Figure 2b).

### Correlative Tissue Analysis

The distribution and quantification of ASMs in brain tissue was assessed by MALDI MSI and LC-MS (Supplementary Table 1). There were no significant relationships between levetiracetam or perampanel tissue concentration and HFO rates at adjacent electrode contacts (Figure 1c-d). Levetiracetam tissue concentrations in Cohort 2 were on average 110-fold greater than perampanel tissue concentrations in Cohort 1 by LC-MS (mean 87.5 uM vs 0.80 uM). Levetiracetam signal by MALDI MSI was relatively homogenously distributed across slices whereas perampanel demonstrated microscopic hotspots over a very low background signal level (Figure 3a).

**Figure 3.**
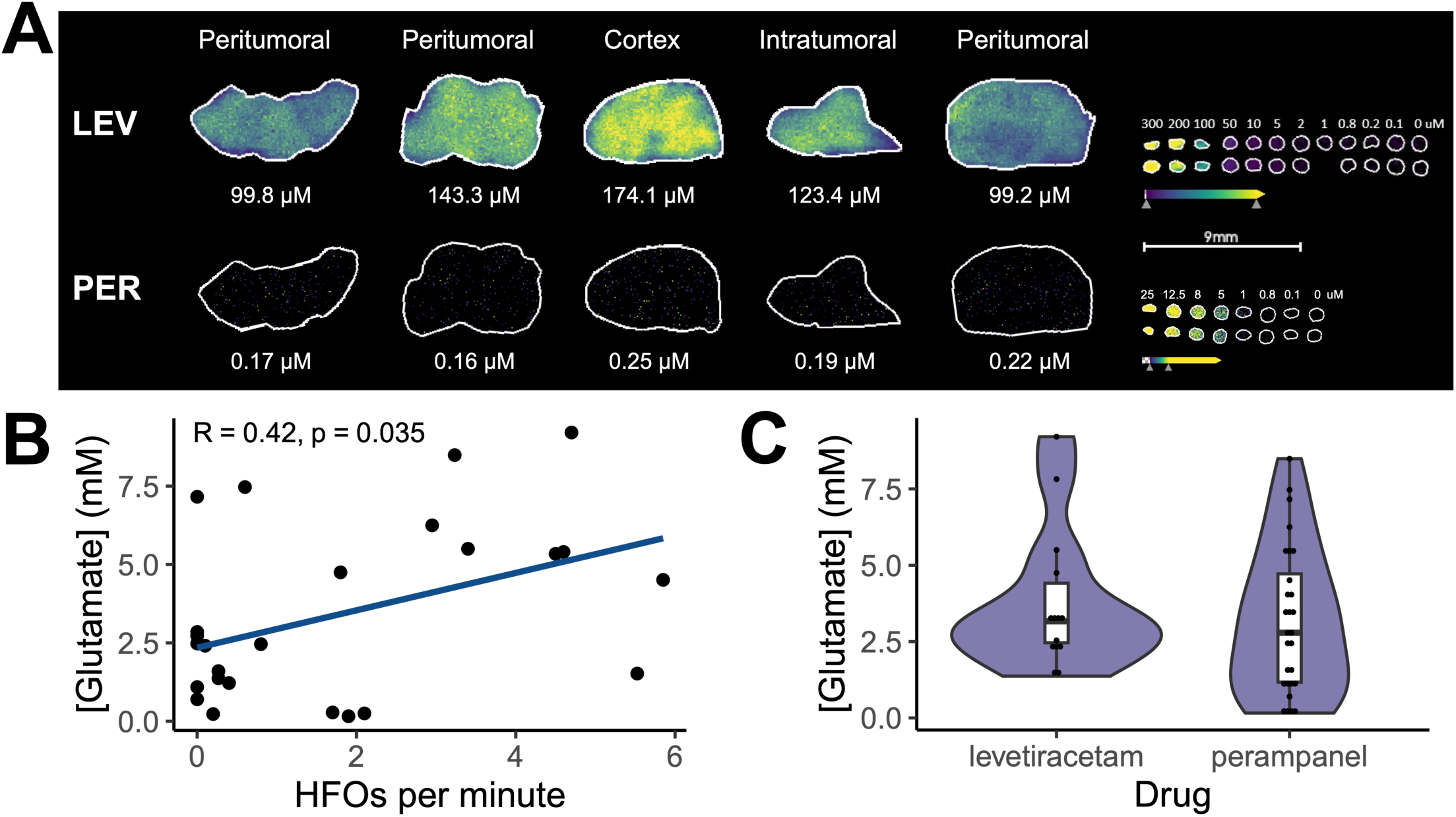
Correlative tissue glutamate analysis. (A) Representative example of MALDI MSI for perampanel and levetiracetam in a patient in Cohort 1 (PER). High residual tissue levetiracetam levels were seen in this case despite a 5 half-life washout period. (B) Positive correlation between tissue glutamate concentration and HFO rate in aggregate (Cohort 1 + Cohort 2). (C) Tissue glutamate concentrations were similar between patients in Cohort 1 (PER) versus Cohort 2 (LEV).

Tissue glutamate concentration and lactate/pyruvate ratio were measured as correlative biomarkers of hyperexcitability and excitotoxicity. Glutamate concentrations in surface biopsies were positively associated with HFO rates in adjacent contacts (Figure 3b) in fixed-effects univariate (β=0.30, SE 0.13, p=0.035) and multivariate (β=0.31, SE 0.14, p=0.043) models controlling for treatment assignment (Cohort 1, β=0.19, SE 0.90, p=0.84) and lactate/pyruvate ratio (β=0.01, SE 0.04, p=0.72). This trend persisted in a mixed-effects model accounting for multiple specimens per patient (β=0.29, SE 0.14, p=0.057).

Including both surface and deep specimens, tissue glutamate concentrations were similar between Cohort 1 and Cohort 2 (median 2.8 vs 3.2 mM, p=0.41, Figure 3c). No significant associations existed between tissue concentrations of glutamate and ASMs by MALDI (perampanel: β=3.16, SE 8.42, p=0.71; levetiracetam: β=-0.03, SE 0.02, p=0.20) or by LC-MS (perampanel: β=0.22, SE 1.01, p=0.83; levetiracetam: β=-0.02, SE 0.02, p=0.27).

## DISCUSSION

In this pilot trial of patients with newly diagnosed HGG, treatment with a targeted AMPA-R antagonist did not demonstrate clinically meaningful effects on peritumoral hyperexcitability or survival. The rapid loading regimen of perampanel for peri-operative seizure control was safe and well-tolerated, similar to the standard approach using levetiracetam. Continued maintenance therapy with perampanel in patients with and without pre-operative seizures was also safe and effective for seizure control, although the study was not powered for this efficacy outcome.

There were no significant differences in peritumoral high frequency activity with targeted post-synaptic AMPA-R blockade by perampanel compared to non-targeted presynaptic modulation of synaptic vesicle release by levetiracetam. Both perampanel and levetiracetam are broad spectrum ASMs and effective clinically for the management of glioma-related epilepsy.^1^ However, it is notable that usual doses of perampanel administered are 2-3 orders of magnitude smaller than doses of levetiracetam (i.e., 4-12mg versus 1000-4000mg per day). We observed a similar difference in brain tissue concentrations of the drugs, with perampanel near the limit of detection for MALDI. This effect was also influenced by averaging pixel intensities over each slice, therefore averaging out small foci with high perampanel concentrations and explaining the differences in perampanel quantitation between MALDI and LC-MS. Nevertheless, it is possible that higher overall brain tissue concentrations of perampanel are necessary to inhibit pathologic synchronized neuronal firing measured by high gamma activity than are needed to raise the clinical seizure threshold.

Several sources of bias may have impacted the effect size seen for perampanel in this trial. Steady state concentrations expected with daily doses of 4mg were likely not achieved at the time of craniotomy despite an aggregate pre-operative load of 10mg. All of the patients with pre-operative seizures were treated with perampanel, while none of those in the standard of care cohort had pre-operative seizures. Thus, the pre-treatment baseline level of hyperexcitability may have been greater in the perampanel treatment cohort. In contrast, levetiracetam was detectable in the perampanel cohort despite a washout period of at least 5 half-lives. This may be due to delayed elimination of levetiracetam from the CNS, and the presence of both drugs would have biased towards a greater decrease in hyperexcitability.

The methodology applied here identified focal peritumoral hyperexcitability with sampling of both active and inactive sites using ECoG grids traversing from tumor to normal appearing cortex. The correlation between tissue glutamate concentrations and HFO rates align with established mechanisms of glioma-related excitability involving increased glutamate release and receptor activation contributing to epileptiform activity.^2,37^ The anatomic correlation with increased HFO rates in the peritumoral margins also reinforce the validity of these signals for glioma-related hyperexcitability.^25,26^ These findings support high frequency activity (i.e., high gamma power or high frequency oscillations) as a promising outcome measure in future surgical window of opportunity trials. We aimed to mitigate some limitations in implementing and interpreting HFO analysis, largely due to variability in methodologies and poor inter-rater agreement,^38^ by using an automated detector with a relatively low threshold to maximize sensitivity, followed by manual visual review to exclude false positive detections.

The presence of glioma-neuron interactions has now been established, however there are likely multiple mechanisms driving synaptogenesis and non-synaptic peritumoral hyperexcitability.^39–41^ Whether treatment aimed at reducing neuronal firing impacts the clinical course of disease, as suggested in animal models and some small retrospective cohorts,^5,12–15,17^ remains uncertain. For example, early glioma-related hyperexcitability in clinical EEG recordings has been associated with decreased overall survival despite treatment with ASMs,^42^ while meta-analysis and a large pooled study of contemporary clinical trials for glioblastoma found no associations between the use of valproic acid or levetiracetam and survival outcomes.^10,16^ In this study, the sample size of patients with newly diagnosed glioblastoma treated prophylactically with perampanel was small, however aligns with a prior negative trial of talampanel in patients with recurrent disease.^18^ Variability in tumor molecular profiles, epilepsy phenotypes, and ASM prescribing practices may partially account for the conflicting reports of anti-tumor effects of ASMs.

Improved biomarkers of hyperexcitability and their systematic integration into clinical trials is necessary.^43^ While clinical seizure reporting is inherently limited,^44^ the optimal methodologies for measuring electrophysiologic hyperexcitability are also uncertain, with substantial variability between labs. Despite the promise of ECoG signal analysis at the time of surgery, this invasive approach remains highly resource intensive and does not allow for tracking of hyperexcitability over time in response to therapy. This poses a challenge to standardizing biomarkers and measuring the impact of treatments targeting neuronal activity over the dynamic course of disease.

In summary, a peri-operative loading regimen of perampanel was safe and well-tolerated, with similar peritumoral hyperexcitability as in levetiracetam-treated patients. Tissue glutamate concentrations correlated with localized high frequency activity. Maintenance anti-glutamatergic therapy was not observed to impact survival outcomes. Further correlative biomarker studies are warranted.

## AUTHORSHIP

Study design: S.T., P.Y.W., N.Y.R.A., J.W.L.; Data acquisition: S.T., M.S.R., M.K.C.D., D.R., E.L., M.A.H., S.S., M.M.M., O.A., W.L.B., E.A.C., A.J.G., M.A.M., T.R.S.; Statistical analysis: S.T., M.S.R., M.A.H.; Data interpretation: S.T., M.S.R., M.A.H., S.S., K.L.L., P.Y.W., N.Y.R.A., J.W.L.; Manuscript preparation: S.T.; Manuscript editing: S.T., M.S.R., M.K.C.D., D.R., E.L., M.A.H., S.S., S.S., M.M.M., O.A., W.L.B, E.A.C., A.G., M.A.M., T.R.S., K.L.L., P.Y.W., N.Y.R.A., J.W.L.

## DATA AVAILABILITY

The datasets generated during and/or analyzed during the current study are available from the corresponding author on reasonable request.

## ACKNOWLEDGEMENTS

The authors would like to acknowledge the Dana-Farber Cancer Institute Oncology Data Retrieval System (OncDRS) for the aggregation, management, and delivery of the clinical and operational research data used in this project. The content is solely the responsibility of the authors. We would also like to thank the patients and families for their participation.

