## Supplementary Data for "Pilot trial of perampanel on peritumoral hyperexcitability and clinical outcomes in newly diagnosed high-grade glioma"

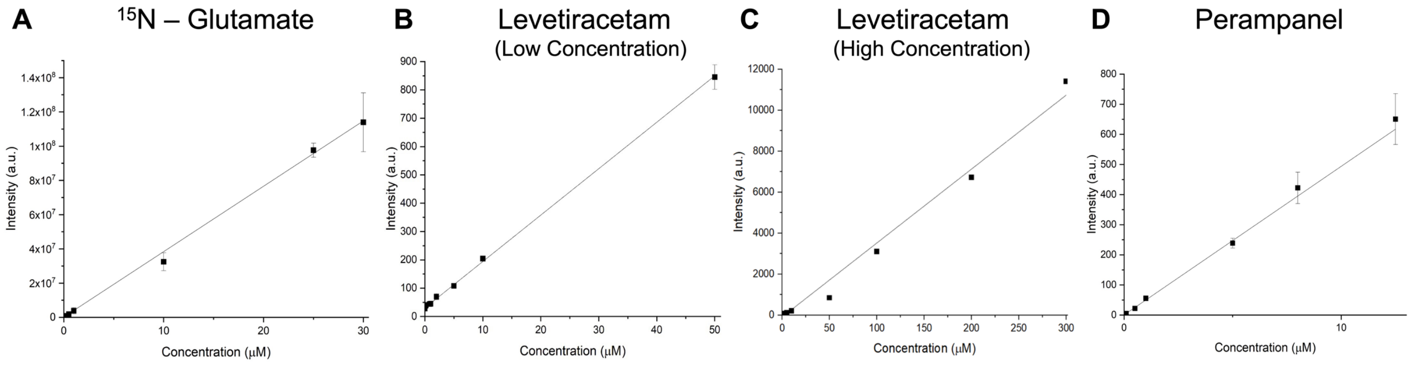

**Supplementary Figure 1.** MALDI Calibration Curves. (A) $^{15}N_1$ – glutamate, normalized to internal standard (IS) $D_5$ - glutamate. (B,C) Levetiracetam; low concentration (B) and high concentration (C), normalized to IS $D_6$ – levetiracetam. (D) Perampanel, normalized to IS panobinostat. All calibration curves were calculated using a weighted linear regression ($1/x^2$).

| | | | | | | |
|---|---|---|---|---|---|---|
| **LLOQ** | 0.71 µM | 0.0025 µM | 0.12 µM | 0.00025 µM | 0.0005 µM | 0.00025 µM |
| **LOD** | 0.21 µM | 0.0025 µM | 0.03 µM | 0.00025 µM | 0.0017 µM | 0.00025 µM |

| | **Levetiracetam Conc. (µM)** | | **Perampanel Conc. (µM)** | | **Glutamate Conc. (mM)** | | **Lactate/Pyruvate Ratios** |
|---|---|---|---|---|---|---|---|
| **Patient ID** | **MALDI-ToF** | **LC-MS/MS** | **MALDI-ToF** | **LC-MS/MS** | **MALDI-FTICR** | **LC-MS/MS** | **MALDI-FTICR** |
| Case 1 sample 1a | 99.78 | 172.00 | 0.17 | 1.43 | 4.51 | 5.68 | 19.1 |
| Case 1 sample 3c | 143.28 | 178.00 | 0.16 | 1.44 | 4.06 | 7.48 | 22.7 |
| Case 1 sample 4d | 174.11 | 165.00 | 0.25 | 0.87 | 1.52 | 3.36 | 23.2 |
| Case 1 sample 5e | 123.38 | 151.00 | 0.19 | 1.51 | 1.62 | 5.04 | 17.9 |
| Case 1 sample 6f | 99.18 | 160.00 | 0.22 | 0.86 | 0.70 | 1.58 | 19.1 |
| Case 2 sample 1a | 2.65 | 1.95 | 0.17 | 0.25 | 3.81 | 5.35 | 3.6 |
| Case 2 sample 2b | 2.47 | 5.65 | 0.17 | 0.20 | 4.77 | 1.44 | 2.8 |
| Case 2 sample 3c | 1.48 | 3.61 | 0.17 | 0.10 | 3.86 | 1.49 | 5.1 |
| Case 2 sample 4d | 0.37 | 0.19 | 0.24 | 0.62 | 3.06 | 4.61 | 4.6 |
| Case 3 sample 1a | 43.99 | 56.17 | NA | NA | 9.21 | 9.90 | 6.4 |
| Case 3 sample 2b | 27.22 | 45.80 | NA | NA | 7.82 | 4.00 | 8.4 |
| Case 3 sample 3c | 29.31 | 11.90 | NA | NA | 3.19 | 6.24 | 2.5 |
| Case 3 sample 4d | 66.55 | 88.20 | NA | NA | 3.40 | 6.78 | 11 |
| Case 4 sample 1a | 7.68 | 6.56 | 0.20 | 0.00 | 1.22 | 0.66 | 14.7 |
| Case 4 sample 2b | 7.12 | 9.60 | 0.20 | 0.00 | 2.41 | 3.07 | 13.2 |
| Case 4 sample 3c | 6.20 | 6.01 | 0.18 | 0.00 | 0.23 | 4.09 | 6.3 |
| Case 5 sample 1a | 40.95 | NA | NA | NA | 1.37 | NA | 8.6 |
| Case 5 sample 2b | 39.92 | 12.20 | NA | NA | 1.60 | 0.69 | 7.8 |
| Case 5 sample 3c | 99.00 | 149.00 | NA | NA | 2.46 | 1.09 | 19 |
| Case 6 sample 1a | 2.84 | 1.05 | 0.04 | 0.03 | 0.16 | 4.48 | 1 |
| Case 6 sample 2b | 2.36 | 2.45 | 0.04 | 0.00 | 0.25 | 4.51 | 1.3 |
| Case 6 sample 4d | 1.05 | 0.36 | 0.00 | 0.00 | 0.28 | 1.31 | 0.9 |
| Case 6 sample Frz | 9.11 | 13.10 | 0.10 | 0.23 | 1.02 | 0.38 | 1.7 |
| Case 7 sample 1a | 22.73 | 21.90 | 0.12 | 0.00 | 8.49 | 8.80 | 2.2 |
| Case 7 sample 2b | 39.57 | 27.30 | 0.16 | 0.00 | 5.34 | 6.15 | 4.8 |
| Case 7 sample 3c | 30.07 | 23.60 | 0.11 | 0.00 | 7.47 | 11.90 | 3.3 |
| Case 7 sample 4d | 15.40 | NA | 0.07 | NA | 5.40 | NA | 17 |
| Case 8 sample 1a | 71.39 | 94.50 | NA | NA | 3.14 | 0.77 | 15.5 |
| Case 8 sample 3c | 76.04 | 126.00 | NA | NA | 2.47 | 4.04 | 11.9 |
| Case 8 sample 4d | 54.88 | 129.46 | NA | NA | 5.50 | 1.13 | 10.4 |
| Case 8 sample 5e | 62.14 | 104.23 | NA | NA | 2.54 | 2.93 | 6.8 |

| | | | | | | | |
|---|---|---|---|---|---|---|---|
| Case 8 sample 6f | 67.29 | 116.00 | NA | NA | 4.75 | 4.64 | 14.8 |
| Case 9 sample 1a | 7.14 | 15.10 | 0.16 | 0.85 | 7.16 | 4.96 | 11.6 |
| Case 9 sample 2b | 1.71 | 6.70 | 0.14 | 1.01 | 2.85 | 6.32 | 3.4 |
| Case 9 sample 3c | 0.00 | 1.06 | 0.12 | 0.47 | 3.54 | 2.72 | 3.6 |
| Case 9 sample 4d | 0.40 | 2.40 | 0.15 | 0.76 | 4.02 | 3.38 | 3.3 |
| Case 9 sample 5e | 3.00 | 9.62 | 0.14 | 1.06 | 5.60 | 4.48 | 6.1 |
| Case 10 sample 1a | 0.00 | 4.65 | 0.13 | 0.74 | 6.25 | 5.64 | 24.8 |
| Case 10 sample 2b | 0.00 | 0.33 | 0.13 | 0.56 | 1.20 | 3.02 | 7.6 |
| Case 10 sample 3c | 0.00 | 0.11 | 0.13 | 0.25 | 3.39 | 1.78 | 10.7 |
| Case 10 sample 4d | 0.44 | 0.22 | 0.16 | 0.81 | 3.54 | 3.10 | NA |
| Case 11 sample 1a | 0.00 | 0.14 | 0.09 | 0.79 | 1.09 | 1.07 | 1.9 |
| Case 11 sample 2b | 2.42 | 0.03 | 0.11 | 0.23 | 2.48 | 2.24 | 44.8 |
| Case 11 sample 3c | 0.00 | 0.00 | 0.12 | 1.26 | 2.73 | 1.92 | 5 |
| Case 12 sample 1a | 63.24 | 111.00 | NA | NA | 3.39 | 1.33 | 3.7 |
| Case 12 sample 2b | 142.24 | 92.54 | NA | NA | 2.20 | 4.96 | 4.4 |

**Supplementary Table 1.** Mass Spectrometry Data. Cells with NA indicate exclusion due to treatment assignment or insufficient tissue for analysis. LLOQ, lower limit of quantitation; LOD, limit of detection.
